# Determinants of Anticipated Video-Supported Treatment Acceptability in Drug-Susceptible Pulmonary Tuberculosis

**DOI:** 10.64898/2026.09.10.26362803

**Authors:** Javier A. Lama, Carlos Seas, Jesus Peinado, Marco Tovar, Javier R. Lama

## Abstract

**Setting:** Twenty-three urban healthcare facilities across five geographic jurisdictions of Metropolitan Lima, Peru.

**Objective:** To identify determinants of anticipated video-supported treatment (VST) acceptability among persons with drug-susceptible pulmonary tuberculosis.

**Design:** Observational, cross-sectional study (August–December 2025). Anticipated acceptability was assessed across two domains: acceptability of use (desire to use VST) and willingness to comply (confidence in fulfilling VST requirements). Adjusted prevalence ratios (aPR) with 95% confidence intervals (95%CI) were estimated using robust Poisson regression with Liang-Zeger cluster-robust standard errors.

**Results:** Among 473 participants (median age 35 years; 51.5% male), 61.3% (95%CI: 57.0%–65.9%) reported acceptability of use and 42.3% (95%CI 36.2%–49.4%) willingness to comply with VST. Educational attainment was the strongest predictor of both outcomes. The ability to record and send videos using a mobile phone and continuity of technology access were associated with acceptability of use and willingness to comply with VST, respectively. Diabetes mellitus, illicit drug use, and intention to abandon treatment were associated with reduced willingness to comply with VST.

**Conclusions:** VST acceptability is shaped by educational attainment, digital competency, structural conditions, and clinical and behavioural profile. VST implementation should prioritise digital skills training and incorporate pre-enrolment screening for clinical vulnerability.

## INTRODUCTION

Tuberculosis (TB) remains the leading cause of death from a bacterial pathogen worldwide, with 10.7 million incident cases and 1.23 million deaths in 2024^1^. WHO-recommended standard treatment for drug-susceptible pulmonary TB requires six months of multi-drug therapy under daily treatment support^2^. Health facility-based treatment support (HFTS) — the most widely implemented monitoring strategy — mandates daily attendance at a health facility, imposing logistical, economic, and psychosocial burdens that compromise adherence and treatment outcomes^3–5^. Video-supported treatment (VST, formerly “video directly observed therapy”), in which patients record and transmit a video of each dose for remote supervision, has emerged as an alternative^2, 6^. Randomised trials demonstrated that VST achieves non-inferior treatment outcomes compared with HFTS, while reducing patient burden^7–12^.

Patient acceptability is a prerequisite for sustainable implementation of health interventions^13^. Despite the clinical evidence supporting VST, data on patient acceptability and its independent determinants are limited. Available evidence^14, 15^ has assessed overall preference without distinguishing between two clinically distinct acceptability dimensions: the desire to use VST and the confidence in complying with its requirements. This distinction has direct operational implications — patients who wish to use VST may lack the self-perceived capacity to sustain it, resulting in avoidable non-adherence.

We conducted a cross-sectional study to evaluate anticipated VST acceptability among persons with drug-susceptible pulmonary TB in Peru. Using the Theoretical Framework of Acceptability^16^, we prespecified two primary outcomes: acceptability of use (the degree to which patients would like to use VST) and willingness to comply (the degree to which patients felt confident they could fulfil VST requirements). Secondary objectives were to assess the associations of TB-related knowledge and beliefs, and self-perceived social stigma, with both outcomes.

## METHODS

### Study design and participants

We conducted a cross-sectional study from August to December 2025 in 23 urban healthcare facilities across five health jurisdictions of Metropolitan Lima, Peru. Eligible participants were adults (≥18 years) receiving the standard six-month regimen for drug-susceptible TB under HFTS, either with microbiological confirmation of drug susceptibility or based on clinical diagnosis, with at least one week of treatment at the time of enrolment. Persons with confirmed or suspected extrapulmonary TB or drug-resistant TB were excluded, as their treatment experience, regimen characteristics, clinical course and monitoring requirements differ substantively from drug-susceptible TB, according to local guidelines. Persons receiving home- or community-based treatment support were similarly excluded, as they do not face the same logistical and psychosocial burdens as HFTS. Persons living with HIV were eligible. We used two-stage cluster sampling. In stage one, from 95 eligible public healthcare facilities — defined as those having treated ≥25 drug-susceptible pulmonary TB cases in 2024 by Peruvian Ministry of Health registries — 19 facilities were selected by simple randomisation, stratified proportionally by geographical jurisdiction. During fieldwork, four selected facilities had fewer eligible patients than anticipated; replacement facilities from the same jurisdictions were added, yielding 23 facilities in total. In stage two, up to 25 consecutive eligible participants per facility were recruited using convenience sampling, in order of presentation for treatment support (**Figure 1**). Sample size was calculated assuming 60% acceptability prevalence, 5% precision (α = 0.05), an intra-cluster correlation coefficient of 0.01, yielding a target sample of 475 participants.

**Figure 1:**
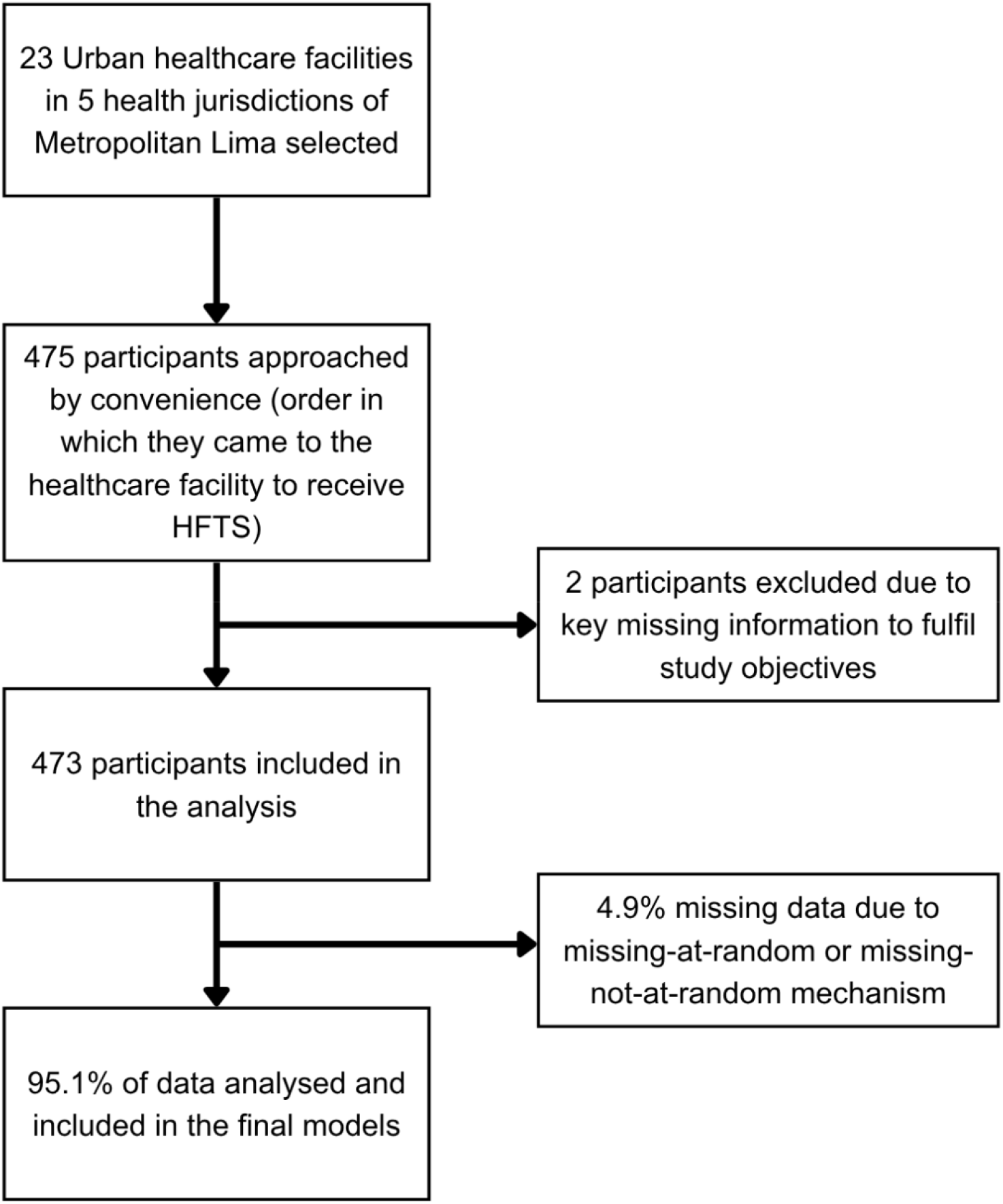
Participant flow diagram.

### Variable definitions and data collection

The primary outcomes correspond to the two domains of the Theoretical Framework of Acceptability^16^ most relevant to implementation: affective attitude and self-efficacy. Acceptability of use (affective attitude) was defined as the degree to which a patient would like to use VST; willingness to comply (self-efficacy) was defined as the degree to which a patient felt confident they could fulfil VST requirements. Both outcomes were assessed using a Likert scale (1–5; higher scores indicating greater acceptability) and dichotomised as negative (1–3) or positive (4–5), allowing a consistent classification suitable for implementation research. The Likert score “3” was classified as negative to ensure conservative interpretation of ambiguous responses and to avoid inflating acceptability prevalence. Independent variables included sociodemographic characteristics, TB-related knowledge and beliefs, structural barriers and facilitators to treatment support, technology access and digital literacy, and self-perceived social stigma, assessed via the validated TB-related Stigma Scale^17^. Data were collected using a structured self-administered paper-based survey and a review of participants’ medical records. The survey was developed following a thorough review of the literature, previous acceptability instruments, and TB-specific data collection tools. Validation followed a three-step framework: content validity was assessed by three experts in TB operational research; face validity was assessed through comprehension testing with five members of the target population; and internal consistency was assessed through pilot testing (n = 48) and calculation of Cronbach’s alpha (= 0.817), indicating high internal consistency. Medical records provided information on TB diagnosis, treatment adherence, and HIV status. Surveys were conducted in Spanish and self-administered without interviewer intervention to minimise social desirability bias.

### Statistical methods

Categorical variables are described as frequencies and proportions; continuous variables as medians and interquartile ranges. Bivariate associations were assessed using the chi-square test for categorical variables, and the Mann-Whitney U test for continuous variables. To identify independent determinants of each primary outcome, we estimated adjusted prevalence ratios (aPR) with 95% confidence intervals (95% CI) using robust Poisson regression with a log link. Because participants were clustered within healthcare facilities — sharing unmeasured sociodemographic, environmental, and structural characteristics — standard errors were adjusted using the Liang-Zeger cluster-robust sandwich estimator, with clustering at the facility level, to prevent artificially narrow confidence intervals^18^. Age, sex, and health jurisdiction were prespecified adjustment variables. Variables with ≥30% missing-not-at-random (MNAR) data were evaluated descriptively only. For variables collected conditionally on a filter question, missing data were recoded to “no”. No variables required exclusion due to collinearity, with a maximum adjusted variance inflation factor of 2.3. Candidate variables for model entry were those with a bivariate P < 0.20 or epidemiological plausibility. For final modelling assessment, backward elimination was conducted using a P ≥ 0.05 threshold based exclusively on cluster-robust Wald test P values. A sensitivity analysis compared the primary model against an alternative model without cluster adjustment; discordant variables are reported as borderline findings. All analyses were performed in R version 4.5.2 (R Core Team).

### Ethics

This study was approved by the Institutional Research Ethics Committee of the Cayetano Heredia Peruvian University and by the ethics committees of all five health jurisdictions of Metropolitan Lima and was classified as minimal risk. Written informed consent was obtained from all participants before enrolment. Anonymity and data confidentiality were guaranteed throughout all stages of data management.

## RESULTS

Of 475 persons enrolled, 473 were included in the analysis; two participants were excluded for not completing the primary outcome items. Participant characteristics are summarised in **Table 1**. The median age was 35 (range: 18–91) years; 51.5% were male. Secondary school was the most frequently reported highest educational level attained (45.1%), and the most common occupation was self-employment (40.8%). Diabetes mellitus was the most prevalent comorbidity (19.0%); 13.1% reported illicit drug use in the past year. At enrolment, 23.7% were in the first month of treatment, 30.7% had documented irregular treatment adherence, and 70.6% had microbiological confirmation of drug susceptibility. HIV co-infection was uncommon (3.4%).

**Table 1:** Participant characteristics.

| Characteristic | Frequency (N = 473)<sup>1</sup> |
| --- | --- |
| Age (years) | 35 [25, 47] |
| Male sex | 243 (51.5) |
| Highest educational degree obtained |  |
| Primary school | 63 (13.5) |
| Secondary school | 211 (45.1) |
| Technical education | 110 (23.5) |
| College education | 84 (17.9) |
| Currently studying | 85 (18.0) |
| Diagnosis of diabetes mellitus | 90 (19.0) |
| Illicit drug or substance use in the past year | 62 (13.1) |
| Month of anti-TB treatment |  |
| 1–2 | 200 (42.4) |
| 3–6 | 272 (57.6) |
| Irregular treatment adherence<sup>2</sup> | 145 (30.7) |
| HIV co-infection<sup>3</sup> | 16 (3.4) |

<sup>1</sup> For categorical variables, absolute count and percentage between parenthesis are presented. For continuous variables, median and quartiles between brackets are presented.
<sup>2</sup> Defined as having missed $\geq 3$ doses in the first two months of treatment, or $\geq 5$ consecutive or cumulative doses throughout the entire treatment duration, as defined by the Peruvian Ministry of Health.
<sup>3</sup> HIV: human immunodeficiency virus.

### Acceptability of use

Overall, 290 participants (61.3%; 95% CI 57.0%–65.9%) reported positive acceptability of use. Educational attainment was the strongest predictor, showing a stepwise positive association: secondary school (aPR 1.98, 95% CI 1.36–2.89), technical education (aPR 2.16, 95% CI 1.44–3.25), and college education (aPR 2.08, 95% CI 1.39–3.12), each relative to primary school. The ability to record and send videos using a mobile phone (aPR 2.04, 95% CI 1.53–2.73), having a safe and private space at home for medication storage (aPR 1.64, 95% CI 1.19–2.25) and greater disruption of daily routine by current HFTS (aPR 1.28, 95% CI 1.08–1.51) were also independently associated with acceptability of use. Conversely, having considered abandoning TB treatment was independently associated with lower acceptability of use (aPR 0.59, 95% CI 0.40–0.86) (**Table 2**).

**Table 2:**
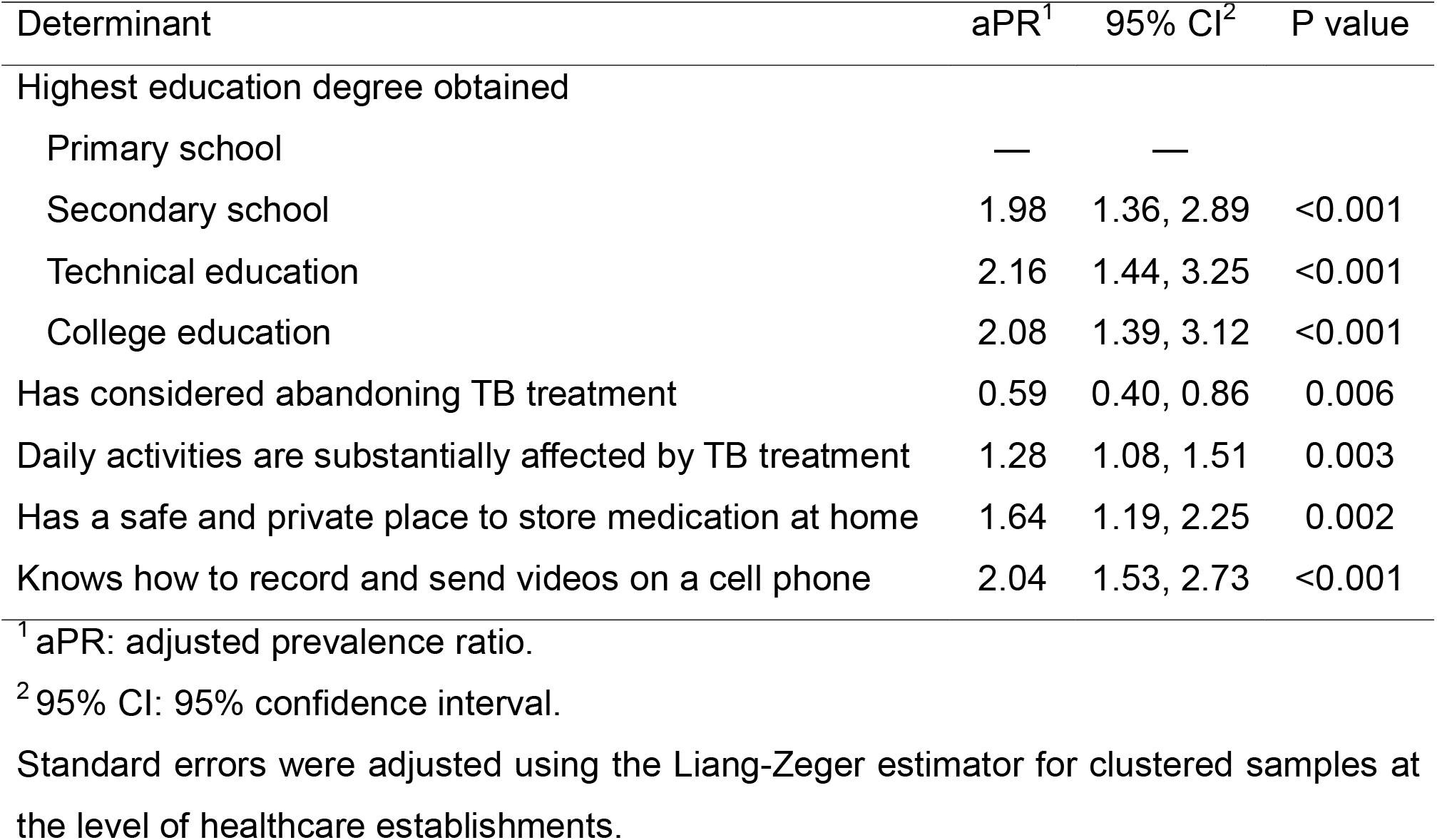
Determinants of acceptability of use.

### Willingness to comply

Only 200 participants (42.3%; 95% CI 36.2%–49.4%) had willingness to comply with VST requirements. Educational attainment again showed a stepwise positive association: secondary school (aPR 1.58, 95% CI 1.03–2.42), technical education (aPR 2.15, 95% CI 1.27–3.65), and college education (aPR 1.76, 95% CI 1.12–2.76). Greater routine disruption due to HFTS (aPR 1.45, 95% CI 1.14–1.83) and having a safe space for medication storage at home (aPR 1.75, 95% CI 1.10–2.76) replicated the pattern observed in the first model. Anticipated access to a capable cell phone in the next six months increased willingness to comply (aPR 1.45, 95% CI 1.16–1.81), while loss of mobile internet access in the past six months reduced it (aPR 0.76, 95% CI 0.62–0.93). Four clinical and behavioural factors were independently associated with lower willingness to comply: diabetes mellitus (aPR 0.59, 95% CI 0.39–0.91), illicit drug use in the past year (aPR 0.64, 95% CI 0.44–0.92), difficulty adhering to current HFTS requirements (aPR 0.66, 95% CI 0.45–0.96), and having considered abandoning TB treatment (aPR 0.65, 95% CI 0.43–0.98) (**Table 3**).

**Table 3:** Determinants of willingness to comply.

| Determinant | aPR<sup>1</sup> | 95% CI<sup>2</sup> | P value |
| --- | --- | --- | --- |
| Highest degree obtained |  |  |  |
| Primary school | — | — |  |
| Secondary school | 1.58 | 1.03, 2.42 | 0.035 |
| Technical education | 2.15 | 1.27, 3.65 | 0.004 |
| College education | 1.76 | 1.12, 2.76 | 0.014 |
| Has diabetes mellitus | 0.59 | 0.39, 0.91 | 0.016 |
| Recreational or illicit drugs use in the last year | 0.64 | 0.44, 0.92 | 0.017 |
| Has considered abandoning TB treatment | 0.65 | 0.43, 0.98 | 0.040 |
| Daily activities are substantially affected by TB treatment | 1.45 | 1.14, 1.83 | 0.002 |
| Has difficulties to comply with TB treatment | 0.66 | 0.45, 0.96 | 0.030 |
| Has a safe and private place to store medication at home | 1.75 | 1.10, 2.76 | 0.017 |
| Cell phone has lost internet access in the last 6 months | 0.76 | 0.62, 0.93 | 0.009 |
| Will have access to a cell phone with the technology to comply with VST within the next 6 months | 1.45 | 1.16, 1.81 | 0.001 |

<sup>1</sup> aPR: adjusted prevalence ratio.<sup>2</sup> 95% CI: 95% confidence interval.
Standard errors were adjusted using the Liang-Zeger estimator for clustered samples at the level of healthcare establishments.

### TB-related knowledge and beliefs, social stigma, and sensitivity analysis

TB-related knowledge and beliefs were high (median of 69.2% correct answers), and self-perceived social stigma was moderate (52.2% of participants perceived social stigma). These variables showed multiple statistically significant bivariate associations with both acceptability outcomes. Neither domain was retained in the multivariable models, indicating that their bivariate associations are largely explained by educational attainment and structural variables also present in the models. The sensitivity analysis comparing cluster-robust standard errors against a model without cluster adjustment yielded a mean standard error ratio of 0.97 for the acceptability of use model and 1.03 for the willingness to comply model, indicating modest clustering effects. Two borderline discordant variables were identified in the willingness to comply model: secondary school education (P = 0.035 cluster-robust vs. P = 0.082 unadjusted) and treatment abandonment ideation (P = 0.040 vs. P = 0.056). All remaining variables were concordant across both estimators.

## DISCUSSION

### Main finding

This study identifies the independent determinants of anticipated VST acceptability among 473 persons with drug-susceptible pulmonary TB across five health jurisdictions of Metropolitan Lima. Two clinically distinct dimensions of acceptability were prespecified: acceptability of use (61.3% positive) and willingness to comply (42.3% positive). The 19-percentage-point gap between them is a clinically important finding in itself — a substantial proportion of patients who would like to use VST do not feel confident they could sustain its requirements. Educational attainment was the strongest and most consistent predictor of both outcomes, with a stepwise gradient reaching twice the probability of positive acceptability among participants with higher education relative to those with primary school only. Critically, functional digital literacy — operationalised as the ability to record and send videos using a mobile phone — predicted acceptability of use independently of device ownership, while anticipated continuity of technology access exclusively predicted willingness to comply. Diabetes mellitus, illicit drug use, and thoughts of discontinuing treatment independently reduced willingness to comply without affecting acceptability of use, delineating a clinically identifiable subgroup at heightened implementation risk.

### Comparison with evidence

The role of educational attainment is consistent with evidence from African cohorts, where formal education is associated with up to three times greater preference for VST over HFTS^19^. Our findings extend this by disaggregating the educational effect across two distinct acceptability constructs (**Figure 2**). Prior studies have associated video-recording ability and digital literacy with VST acceptability^15, 20^, which is consistent with our finding that functional competency to record and transmit videos was associated with higher acceptability of use. The bidirectional effect of current HFTS burden on VST acceptability is a finding not previously described in the VST literature. Patients whose daily routine is substantially disrupted by HFTS showed higher acceptability of use and willingness to comply with VST — consistent with evidence that logistical interference with employment and family responsibilities is one of the primary motivators for seeking alternative treatment delivery modalities^21, 22^. Conversely, patients reporting difficulty adhering to current HFTS requirements showed reduced willingness to comply with VST. This implementation paradox — that patients with the greatest adherence difficulty are simultaneously those least likely to feel confident sustaining VST — reflects a documented phenomenon: repeated non-adherence progressively erodes self-perceived capacity to meet treatment demands^23^. The prevalence of diabetes mellitus in our sample is consistent with reported rates in Latin American and Asian TB cohorts^24, 25^, and its negative association with willingness to comply reflects the cumulative burden of managing two chronic conditions, reducing capacity for additional monitoring^26^. The association between illicit drug use and lower willingness to comply is concordant with evidence documenting reduced treatment adherence in this population^27^. Reduced anticipated acceptability among patients with treatment abandonment ideation reflects insufficient motivational readiness and unestablished perceived need for medication^28^. Motivational interviewing has demonstrated efficacy in advancing such patients toward readiness for treatment engagement^29, 30^. The attenuation of TB knowledge and stigma associations in multivariable models is best explained by confounding through educational attainment and structural variables.

**Figure 2:**
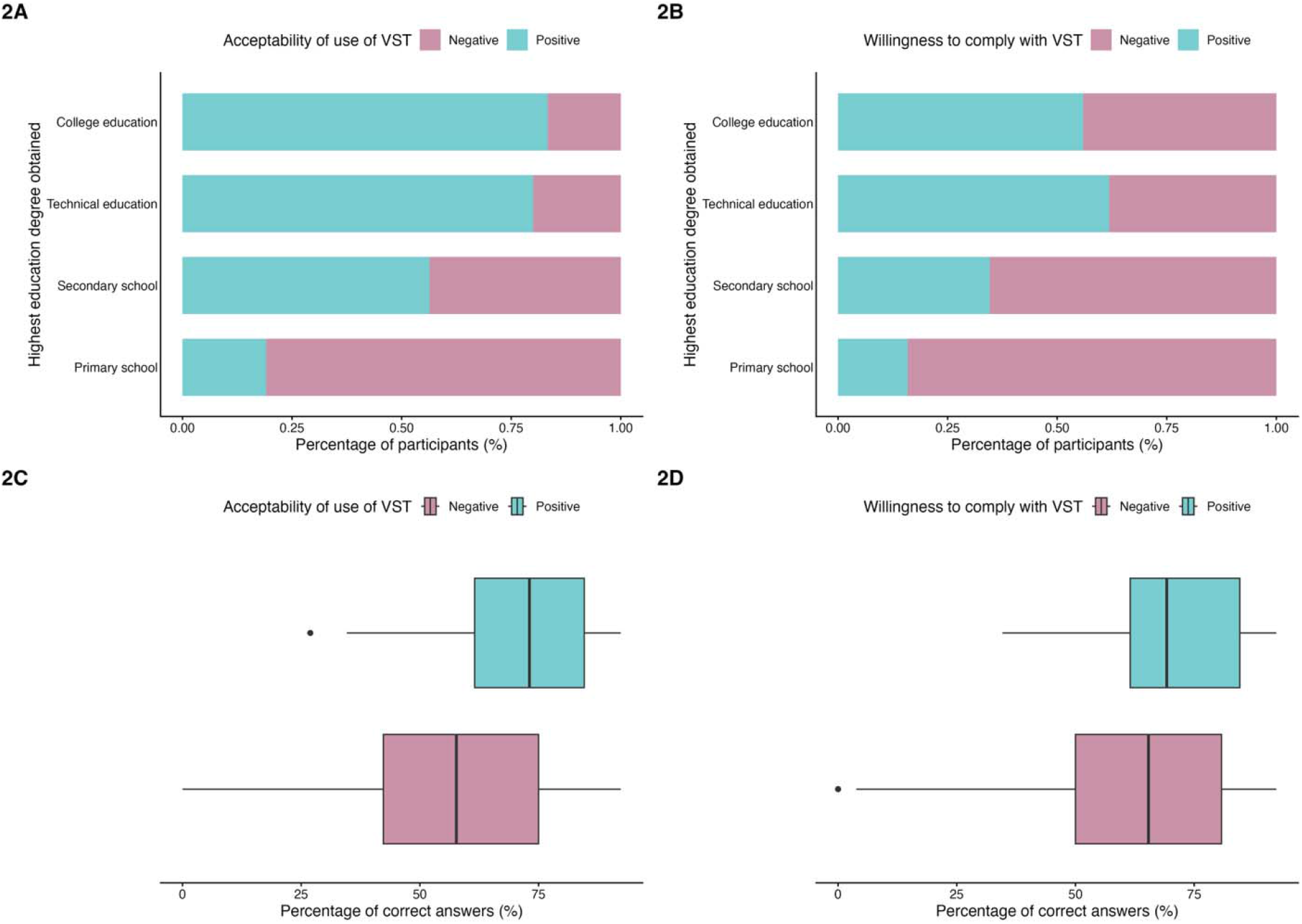
Highest education degree and TB-related knowledge according to VST acceptability. Educational attainment and TB-related knowledge by anticipated VST acceptability. Panels 2A–2B: proportion of participants reporting positive and negative acceptability of use (2A) and willingness to comply (2B), stratified by highest educational level attained. Panels 2C–2D: distribution of TB-related knowledge scores (percentage of correct answers) by acceptability of use (2C) and willingness to comply (2D). TB: tuberculosis; VST: video-supported treatment.

### Clinical and programmatic implications

These findings collectively support a multidimensional pre-enrolment framework for VST implementation, acknowledging that anticipated acceptability is a necessary but insufficient precursor to actual uptake. VST implementation programmes should consider prioritising digital skills training alongside device provision, and screening for vulnerability factors — diabetes mellitus, illicit drug use, and treatment abandonment ideation — that could inform which patients would benefit from preparatory psychosocial support before enrolment. Prospective studies evaluating whether VST implementation preceded by digital literacy training and psychosocial preparation translates anticipated acceptability into improved treatment adherence, clinical outcomes and patient experience represent the logical next step.

### Limitations

Several limitations should be considered. The cross-sectional design precludes causal inference, and results reflect stated intentions rather than observed behavior. The gap between health-related intentions and subsequent behavior^31^ means that anticipated acceptability may not translate into actual VST uptake or adherence. Findings should be interpreted as identifying candidate priorities for implementation research rather than as direct predictors of eventual use. The study excluded persons on home- or community-based treatment support, which limits generalisability to these subgroups. Household income and transport costs were excluded from multivariable models; their effects remain unmeasured. Self-report bias — including social desirability and recall effects — is inherent to survey methodology. Our measure of digital literacy relied on a single behavioural item (video-recording ability) rather than a validated multidimensional instrument, and broader health literacy, prior experience with digital technology, family support, and healthcare worker perspectives on VST were not captured and may explain additional variance in acceptability.

### Conclusion

Anticipated VST acceptability in persons with drug-susceptible pulmonary TB in Metropolitan Lima is a multidimensional phenomenon shaped by educational attainment, digital competency, current treatment burden, structural conditions, and clinical and behavioural profile. Digital literacy predicts acceptability of use independently of device ownership, while continuity of technology access predicts willingness to comply. Potential participants with diabetes mellitus, illicit drug use, or treatment abandonment ideation may benefit from targeted preparatory interventions before VST enrolment. These findings provide an evidence-based framework for equitable VST implementation in high-burden urban settings and establish the rationale for prospective studies of skills-based and psychosocial pre-enrolment interventions.

## Data Availability

All data produced in the present study are available upon reasonable request to the authors.

## ACKNOWLEDGEMENTS

The authors thank the study community health workers and staff of the 23 participating healthcare facilities across the five health jurisdictions of Metropolitan Lima for their support during outreach work and data collection, as well as all participants who generously provided their time to this study. We also thank Ms. Michael W. Louella for his editorial revision and draft manuscript.

## Funding

This study was supported by the “Promoción Medicina 1989” Research Grant awarded by the Universidad Peruana Cayetano Heredia (2023).

## Author contributions

JAL, JP, MT and JRL conceived and designed the study, developed and validated the data collection instrument. JAL coordinated field data collection, performed all statistical analyses, and drafted the manuscript. CS, JP, MT and JRL contributed to critical revision of the manuscript for important intellectual content. All authors read and approved the final manuscript version.

## Conflicts of interest

The authors declare no conflicts of interest. The funding source had no role in study design, data collection, analysis, interpretation, or manuscript preparation.

## Notes

### Competing Interest Statement

The authors have declared no competing interest.

### Author Declarations

Ethics committee/IRB of Universidad Peruana Cayetano Heredia gave ethical approval for this work.

